# A novel SEIR-e model for disease transmission and pathogen exposure

**DOI:** 10.1101/2022.02.16.22271093

**Authors:** Tamara A. Tambyah, Matthew J. Testolin, Alexander M. Hill

## Abstract

In this study, we couple compartment models for indoor air quality and disease transmission to develop a novel SEIR-e model for disease transmission and pathogen exposure. In doing so, we gain insight into the contribution of people–people and people–pathogen interactions to the spread of transmissible diseases. A general modelling framework is used to assess the risk of infection in indoor environments due to people–pathogen interactions via inhalation of viral airborne aerosols, and contact with contaminated surfaces. We couple the indoor environment model with a standard disease transmission model to investigate how both people–people and people–pathogen interactions result in disease transmission. The coupled model is referred to as the SEIR-e model. To demonstrate the applicability of the SEIR-e model and the novel insights it can provide into different exposure pathways, parameter values which describe exposure due to people–people and people–pathogen interactions are inferred using Bayesian techniques and case data relating to the 2020 outbreak of COVID-19 in Victoria (Australia).

## 1 Introduction

Outbreaks of infectious diseases, such as influenza or Ebola, can result in pandemics or epidemics and widespread mortality [1,2]. Mathematical modelling of disease outbreaks is often used to propose and assess the effectiveness of preventative measures, and to quantify the rate of disease transmission [3]. In 2019 a new coronavirus, severe acute respiratory syndrome coronavirus 2, SARS-CoV-2, was found to cause the respiratory illness COVID-19 which has since resulted in a global pandemic with over 5.6 million deaths as of February 3, 2022 [4,5]. Transmission of COVID-19 typically occurs in indoor environments, such as workplaces or households, where infected individuals spend extended periods of time with others [6]. Infection prevention and controls measures, such as masks, social distancing and stay at home orders, have been implemented in some countries to reduce the spread of COVID-19, and more recently vaccination has become available. While studies into the effectiveness of such measures continue to evolve, available evidence suggests that SARS-CoV-2 is spread through close contact with infected individuals via inhalation of airborne respiratory droplets [3,6–8]. Transmission through fomites, such as contaminated surfaces, may occur if fomite exposure is followed by contact with the mouth or nose [3, 6–8]. Some studies indicate that indoor air quality may also affect COVID-19 transmission [8–10].

Compartment models are routinely used to study indoor air quality and disease transmission. For indoor air quality, compartments are used to divide the environment into multiple zones where the air in each compartment is assumed to be well–mixed, and the air exchange rate between compartments is known [11–13]. For disease transmission, compartments are used to distinguish between the number of people who are susceptible to, exposed to, infected with, and recovered from an infectious disease. Such models are referred to as SEIR models, and used to study disease transmission resulting from direct contact between susceptible and infected individuals [14–17]. Some studies include additional compartments to differentiate between symptomatic and asymptomatic infections [3, 18–20], or to investigate how interaction with the environment mediates the spread of disease [21–24]. In this study, we take a different approach and develop a physics–based compartment model of an indoor environment with analytic solutions, referred to as the environment model, to investigate how indoor air quality, inhalation of airborne aerosols and fomite transmission from people–pathogen interactions result in adverse health effects, such as infection. The model is parametrised for SARS-CoV-2 to study the transmission of COVID-19 in indoor environments, where a dose–response curve is used assess health effects based on the duration of exposure and dose received. To further study the spread of transmissible infectious diseases due to people–people and people–pathogen interactions, we develop the SEIR-e model by coupling the environment model with a standard SEIR model, and derive an expression for the basic reproduction number, ℛ_0_.

Due to the direct threat of COVID-19 outbreaks, studies use standard disease transmission models to estimate the incubation, *γ*^−1^, and infectious, *δ*^−1^, periods of COVID-19 [25–28]. Such studies also estimate the contact parameter, *β*, which reflects the rate at which susceptible and infected individuals come into direct contact [25–27]. The basic reproduction number, ℛ_0_ = *βδ*^−1^, measures the average number of secondary infections from a single infected individual, and is often used to assess the effectiveness of infection prevention and control measures [3, 14, 24, 29, 30]. We apply the SEIR-e model to COVID-19 case data, and use Bayesian techniques to estimate the incubation and infectious periods of COVID-19. In addition, we estimate parameters which reflect the rate of transmission due to people–people and people–pathogen interactions to investigate the contribution of different exposure pathways to *β*. While the model is parametrised for SARS-CoV-2, it is also applicable to studying other types of contaminants such as pathogens (SEIR-e model), and chemical or radiological (environment model) materials.

## 2 Model description

In this section, we formulate a physics–based compartment model for indoor air quality, referred to as the *environment model*, to investigate how exposure to pathogens or other contaminants in indoor environments can result in health effects. By coupling the environment model with a standard SEIR model for disease transmission, we develop the *SEIR-e model* to study how people–people and people–pathogen interactions affect the spread of a transmissible disease.

### 2.1 Environment model

The environment model is a compartment model for assessing the health effects of exposure to contaminated material in indoor environments. While the environment model can be applied to any chemical, biological or radiological material, we focus on pathogens in order to study the transmission of SARS-CoV-2 in indoor environments. Two exposure routes are considered: (1) inhalation of viral airborne aerosols, and (2) contact with contaminated surfaces. Compartments are used to represent the concentration of airborne aerosols, the amount of deposited surface contaminant, and the total dose received due to inhalation and contact (Fig 1).

**Figure 1:**
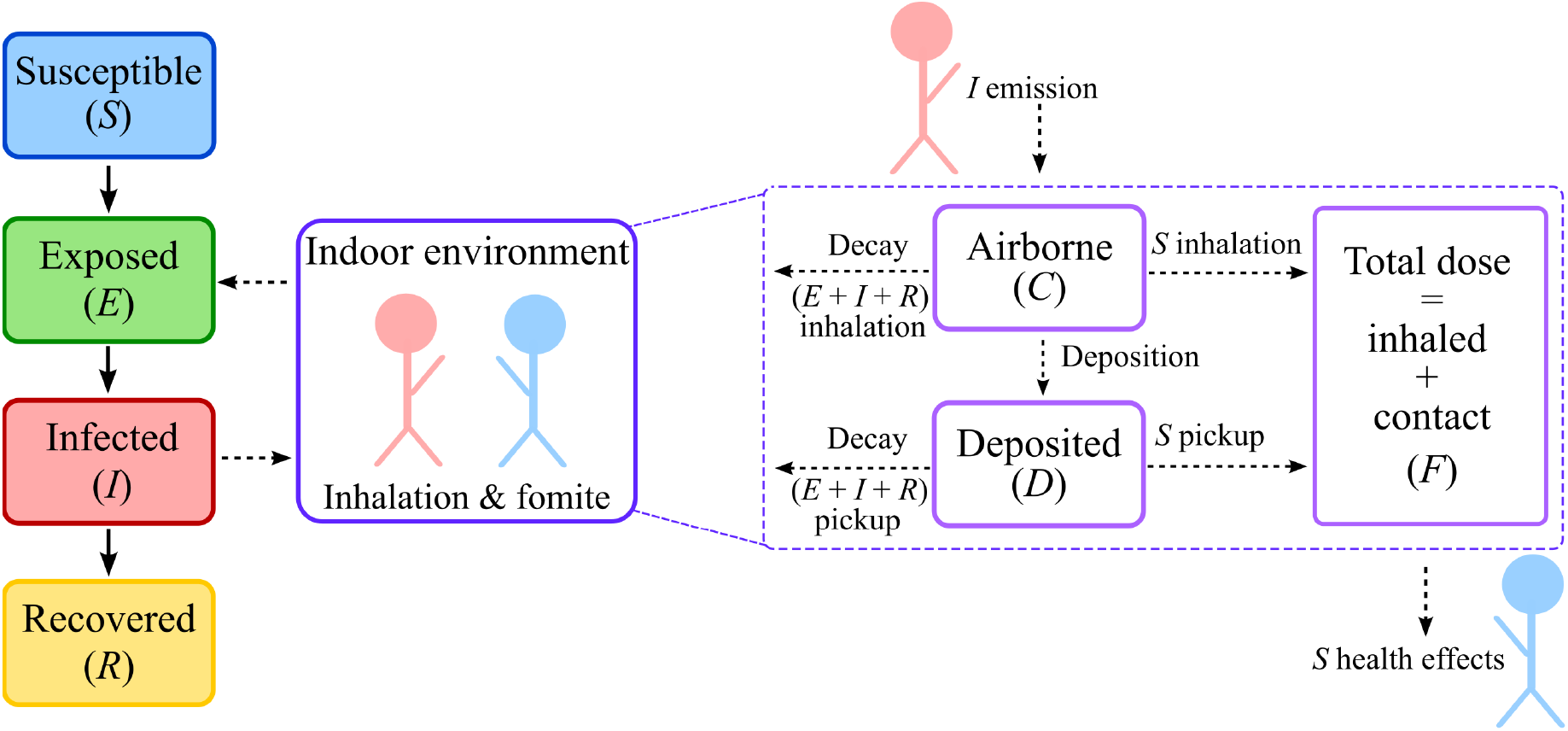
SEIR-e model schematic where the environment model is shown in purple. Compartments represent the number of people who are susceptible to (*S*), exposed to (*E*), infected with (*I*) and recovered from (*R*) the disease, the amount of airborne (*C*) and deposited (*D*) material in the environment, and the total dose received (*F*) by susceptible individuals. The solid and dashed arrows represent the transfer of people and pathogens respectively between compartments. The physical processes included in the environment model are shown in terms of sources and sinks.

Similar to other compartment models for indoor air quality [11,12], the environment model is not spatially resolved. Liquid droplets are categorised by diameter into size bins using a distribution. The distribution and number of size bins, *z*, depends on the type of contaminant, and the mechanism used to release contaminant into the system. We define *C*_*i,t*_, *D*_*i,t*_ and *F*_*i,t*_ for *i* = 1, 2, …, *z* to represent the concentration of airborne material, amount of deposited surface material and total dose of the *i*^th^ size bin at time *t*. As only susceptible individuals are at risk of health effects, we assume that all susceptible individuals receive the same dose via inhalation and surface contact such that *F*_*i,t*_ represents the total dose received by a single susceptible individual. Following [31], we represent physical processes as sources and sinks using iterative statements which describe the solution in the time interval [*t* − Δ*t, t*) for the *i*^th^ size bin:

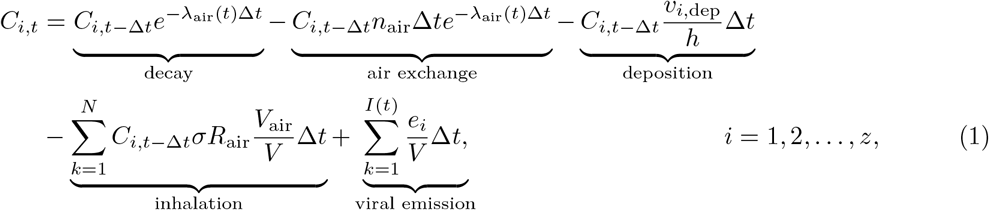

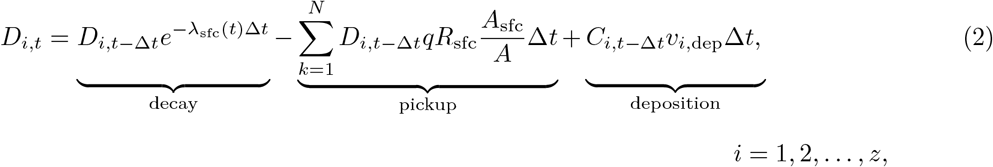

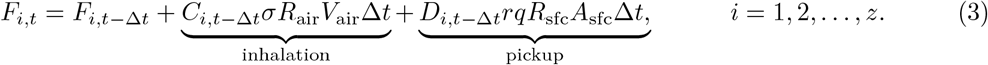

Each term in Eqs (1)–(3) corresponds to a physical process in Fig 1. The viral emission term in Eq (1) represents that, in the context of SARS-CoV-2, viral contaminant is released into the system as an airborne aerosol via infected individuals coughing, speaking and breathing. The bronchial/laryngeal/oral (BLO) model is used to characterise the droplet size distribution, and to determine the emission rate of droplets in each size bin, *e*_*i*_ (Fig 2 (a)). The BLO model and its application to SARS-CoV-2 is outlined in [31]. The contaminant is assumed to be instantaneously well–mixed with ambient air, and the indoor environment is assumed to be well–ventilated such that clean ambient air exchanges with airborne contaminant at rate *n*_air_. These assumptions give rise to the air exchange term in Eq (1). The decay terms in Eqs (1)–(2) account for the degradation and deactivation of airborne and surface contaminant, where the time dependent airborne and surface decay rates, *λ*_air_(*t*) and *λ*_sfc_(*t*) respectively, vary with temperature (*T*) and relative humidity (RH) [32, 33] (Fig 2(b)). Deposition of airborne aerosols onto horizontal surfaces via gravitational settling and diffusion is approximated by the deposition term in Eqs (1)–(2), where the rate of deposition depends on the height of the room, *h*. The deposition velocity, *v*_*i*,dep_, increases with droplet size, and large droplets above a threshold are assumed to deposit instantaneously in order to conserve mass.

**Figure 2:**
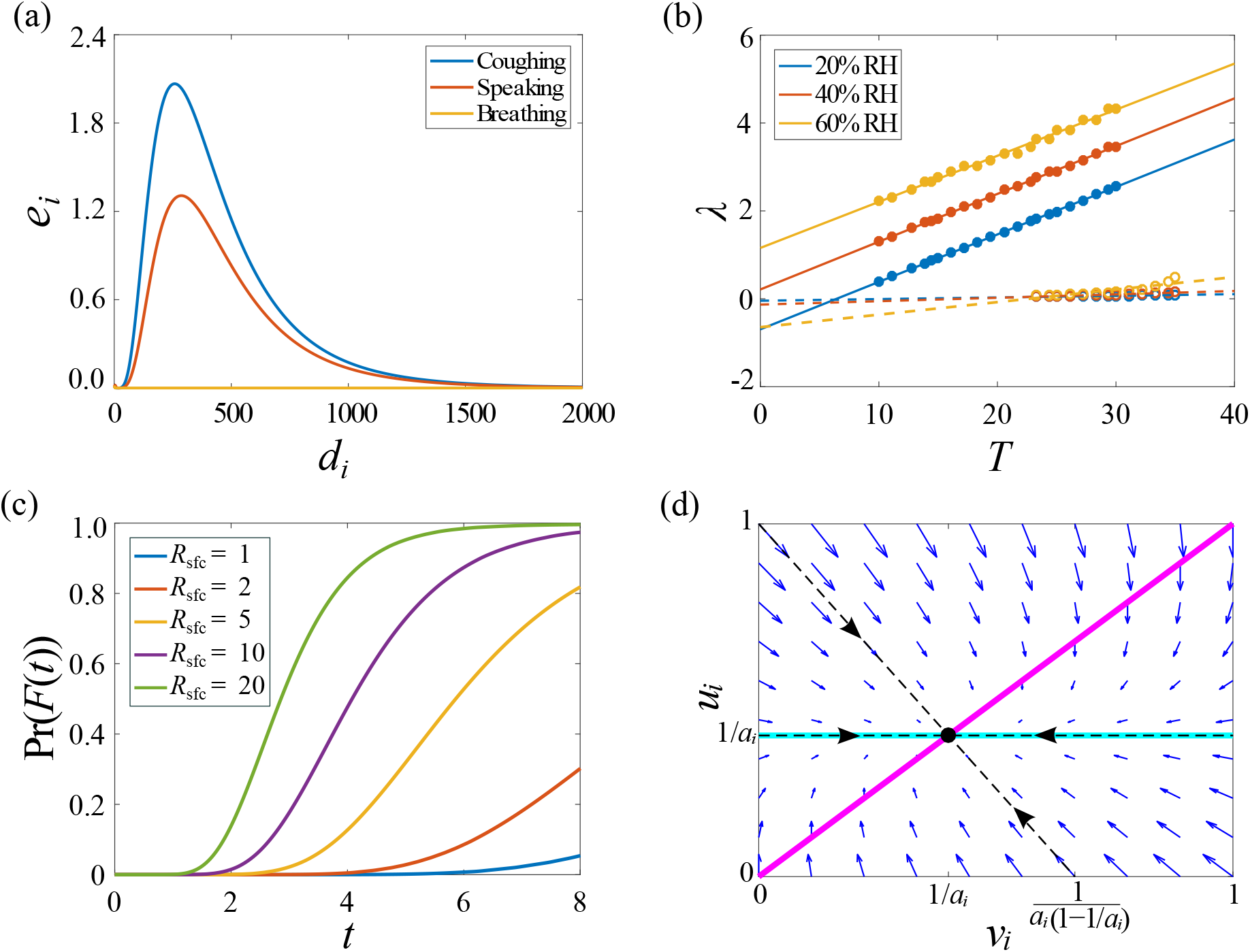
Application of the environment model to SARS-CoV-2. (a) Droplet distribution for SARS-CoV-2 resulting from infected individuals coughing, speaking and breathing (BLO model), where the droplet diameter, *d*_*i*_, is measured in *μ*m and *e*_*i*_ in virus/min. (b) Airborne (solid) and surface (dashed) decay rates for different relative humidity conditions where *λ* is measured in hour^−1^ and *T* in °C. (c) Analytic solutions for varied surface contact rate where the probability of health effects, Pr(*F* (*t*)), is computed using a dose–response curve (Eq (9)) and *t* is the duration of exposure measured in hours. (d) Phase plane for a single size bin showing the fixed point 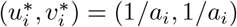 (black dot), *u*_*i*_ nullcline (cyan), *v*_*i*_ nullcline (magenta), eigenvectors (dashed black), eigenvalues (black arrows) and gradient field (blue arrows).

All *N* individuals in the environment are assumed to inhale airborne aerosols, and remove surface contaminant via the hands, referred to as *pickup*. In Eqs (1)–(2), the inhalation and pickup terms are included using summations such that *C*_*i,t*_ and *D*_*i,t*_ represent the total concentration of airborne and deposited material in the environment. Assuming all susceptible individuals receive the same dose, corresponding summations are not required in Eq (3) as *F*_*i,t*_ is the dose received by a single susceptible individual. The inhalation term represents that breathing occurs at a rate *R*_air_ where a fraction of the environment is inhaled with each breath, *V*_air_*/V*. We assume that only droplets within the respirable range are inhaled by choosing *σ* = 1 if *i* ≤ *j* and 0 otherwise, where *j* denotes the number of size bins within the respirable range. As larger droplets deposit quicker than smaller droplets, we choose *j* to reflect the distance from the source at which droplets are assumed to be respirable. For example, small values of *j* correspond to the assumption that only small droplets can be inhaled. That is, exposed individuals are located some distance away from the source. Conversely, large values of *j* relate to a larger respirable range such that exposed individuals are assumed to be located near the source. The pickup term corresponds to a fraction of surface material, *A*_sfc_*/A*, being removed via the hands at a rate *R*_sfc_ with efficiency *q*. We assume that contaminant is transferred from the hands to the mouth or nose at efficiency *r*.

As Eqs (1)–(3) describe the solution at discrete time points, we now derive an analogous system of ordinary differential equations to describe the rate of change of the solution with time. We introduce the field functions

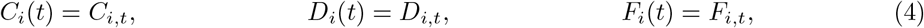

to represent the airborne concentration, deposited material and total dose of the *i*^th^ size bin, where *t* is a continuous time variable. Substituting Eq (4) into Eqs (1)–(3), expanding all terms in a Taylor series about *t*, taking the limit as Δ*t* → 0 and neglecting 𝒪(Δ*t*^2^) gives

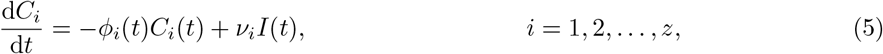

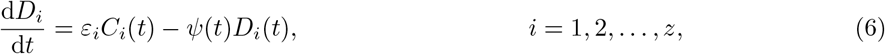

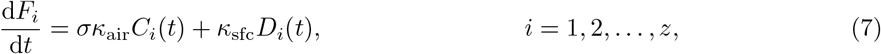

where

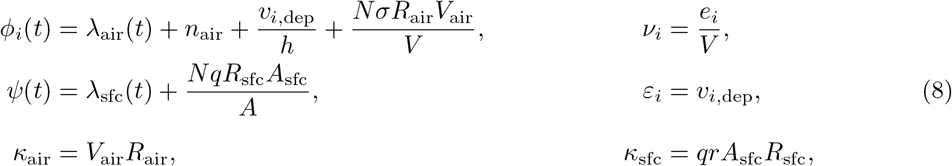

for *i* = 1, 2, …, *z* and *σ* = 1 if 1 ≤ *i* ≤ *j* and 0 otherwise. The environment model is given by Eqs (32)–(34) which is applied to each size bin.

#### 2.1.1 Application to SARS-CoV-2

To apply the environment model to SARS-CoV-2, we consider a scenario, reflective of a workplace environment, where a susceptible individual is exposed to an infected individual in a regulated indoor environment for 8 hours. A probit dose–response curve is used to compute the probability of infection

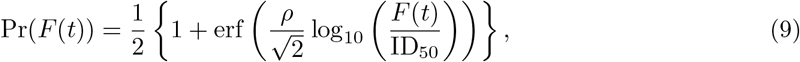

where *F* (*t*) =Σ _*i*_ *F*_*i*_(*t*) is the total dose received due to inhalation and pickup, *ρ* is the probit slope of the dose–response curve, and ID_50_ is the reference dose for 50% probability of infection [31]. Assuming the decay rates are constant in a regulated indoor environment, simple analytical solutions to Eqs (32)–(34) exist for the scenario (*I*(*t*) = 1, *N* = 2). We non-dimensionalise the environment model:

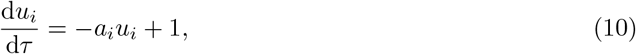

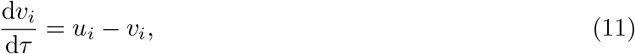

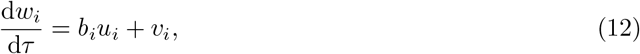

and derive the analytic solutions:

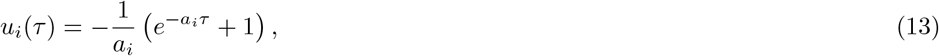

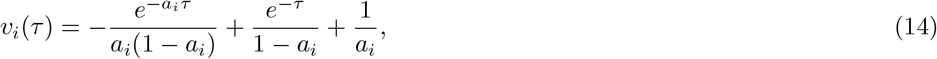

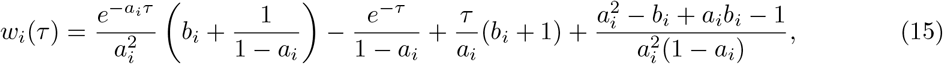

where *u*_*i*_, *v*_*i*_, *w*_*i*_ and *τ* are non-dimensional, *a*_*i*_ =*ϕ*_*i*_*/ψ* and *b*_*i*_ = (*σκ*_air_*ψ*) */* (*κ*_sfc_*ε*_*i*_) (Appendix A.1). Fig 2(c) shows how the probability of COVID-19 infection increases as the contact rate increases. The sensitivity of the environment model to different parameter values relevant to SARS-CoV-2 is further examined in Appendix A.2.

Clearly, Eqs (13)–(15) yield singularities if

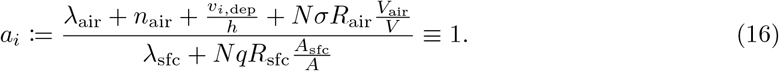

Eq (16) shows that *a*_*i*_ is the ratio of sink processes, such as inhalation and surface removal, and physical decay processes, such as viral decay, deposition and air exchange. As these processes occur on different timescales, we now seek to determine if there exists a dominant process which dictates if *a*_*i*_ ≫ 1 or *a*_*i*_ ≪ 1. Sensitivity analyses in Appendix A.2 show that air exchange, inhalation and surface removal occur on the time scale of hours for *N* = 𝒪(1), as does viral decay (Fig 2(b)). However, the timescale of deposition depends on the droplet size distribution. In the context of SARS-CoV-2, we consider releases where the bulk of material consists of droplets with diameter 200–600 *μ*m and *v*_*i*,dep_ = 𝒪(1) s^−1^ (Fig 2(a)). This implies that deposition is the dominant process such that *a*_*i*_ ≪ 1, and the singularities are not of concern.

Phase plane analysis reveals that the contribution of airborne and deposited material to the total dose is equivalent as *τ* → ∞ (Fig 2(d)). As *τ* → ∞ represents a physical timescale on the order of days, the applicability of the environment model is limited to isolated indoor settings where people are confined for extended periods of time, such as households. In the context of SARS-CoV-2, it is likely that people will start to recover and emit less airborne viral material a number of days after infection onset. Thus, the amount of airborne and surface contaminant will decrease and approach the fixed point, 1*/a*_*i*_ ≪ 1, as *τ* → ∞.

### 2.2 SEIR-e model

In constructing the environment model, we consider how people–pathogen interactions result in infection for susceptible individuals who do not interact with others. When considering the spread of infectious diseases, susceptible individuals are likely interact with infectious individuals and be exposed as a result of people–people interactions also. This motivates us to develop the SEIR-e model by coupling the environment model with a standard SEIR model for disease transmission.

We denote the number of people who are susceptible to, exposed to, infected with and recovered from the disease as *S*(*t*), *E*(*t*), *I*(*t*) and *R*(*t*) respectively. Using a dose–response function to compute the probability of infection due to inhalation and surface pickup gives

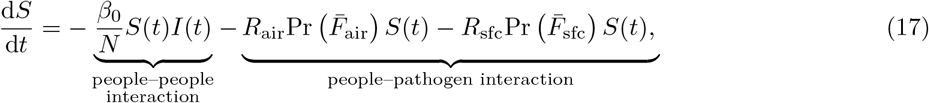

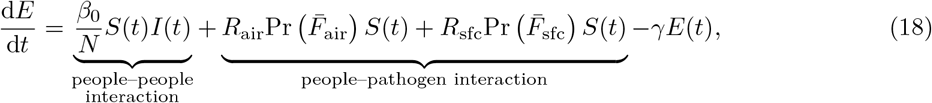

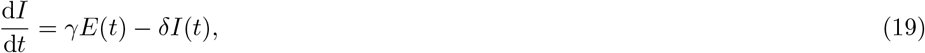

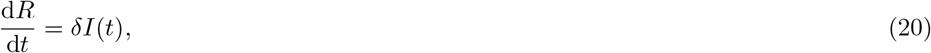

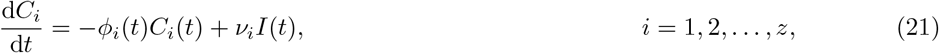

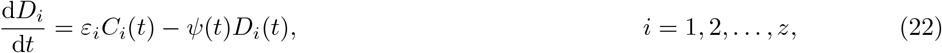

where 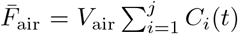 and 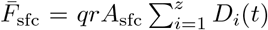 are the instantaneous doses received via inhalation and surface pickup respectively, and *N* = *S*(*t*) + *E*(*t*) + *I*(*t*) + *R*(*t*). The people–people interaction term corresponds to the exposure term in standard SEIR models, where *β*_0_ denotes the progression rate from susceptible to exposed due to contact with infected individuals. However, *β*_0_ ≠ *β* as cited values of *β* may implicitly include transmission due to other exposure pathways. The people–pathogen interaction term accounts for inhalation of airborne aerosols and surface pickup at rates *R*_air_ and *R*_sfc_ respectively. The incubation and infectious periods are *γ*^−1^ and *δ*^−1^ respectively, such that *γ* and *δ* are the progression rates from exposed to infectious, and infectious to recovered. In constructing the SEIR-e model, we assume that all infected individuals continuously emit virus for the duration of their infectious period. That is, *ν*_*i*_ is constant and time independent.

Similar to other disease transmission models [23,24], the Next Generation method is used to derive an exact expression for the basic reproduction number [30]. We let 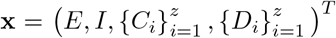 and **y** =(*S, R*) ^*T*^ such that the model can be written as d**x***/*d*t* = ℱ (**x, y**) − *𝒱* (**x, y**) where

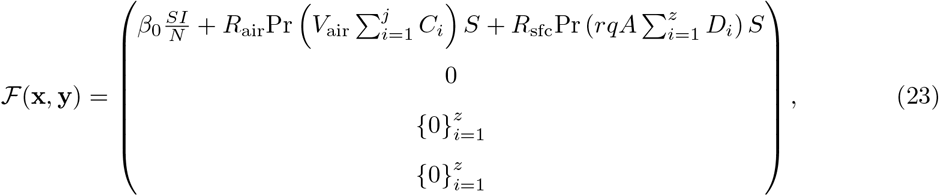

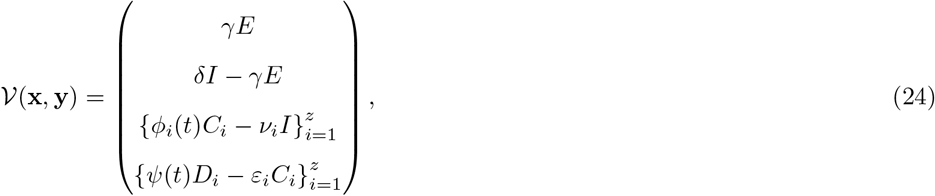

and 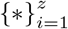 denotes a vector of the expression *\** evaluated from *i* = 1, …, *z*. Computing the spectral radius of the Next Generation Matrix [30] gives

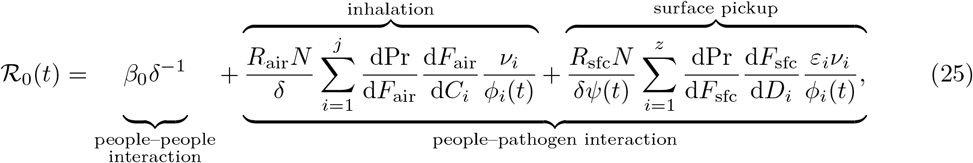

for a fully susceptible population. Eq (25) is an exact expression for ℛ_0_(*t*) that incorporates exposure via inhalation and pickup in addition to contact between susceptible and infectious individuals. A detailed derivation of ℛ_0_(*t*) is provided in Appendix B.

Eqs (17)–(22) and (25) form the SEIR-e model where people–people and people–pathogen interactions are explicitly considered. This distinction is a key feature of the SEIR-e model, and is not included in standard disease transmission models [14–17]. The SEIR-e model can be used to investigate the relative contribution of people–people and people–pathogen interactions to disease transmission in indoor environments such as hospitals, offices, and defence vessels including submarines and aircraft. Forecasting in such environments may also be possible. The SEIR-e could also be used to assess the effectiveness of infection prevention and control measures, such as air handling, cleaning and masks, in indoor environments by modelling the relevant scenarios.

## 3 COVID-19 case study

Parameter values for disease transmission models can be estimated using case data recorded during live outbreaks which reflects the number of people infected. Such estimates are often used to understand the dynamics of outbreaks, and to assist authorities in planning mitigation strategies and caring for the infected [3, 28, 35]. The accuracy of parameter estimates reflects the quality of the data used, and can vary between studies which use different data sets and parameter estimation techniques.

Numerous COVID-19 outbreaks have occurred worldwide since 2019, and recurring outbreaks are common. As such, many studies use disease transmission models and real–time case data to estimate infectious disease parameters relating to COVID-19 [35]. Of the COVID-19 outbreaks that occurred in Australia in 2020, Victoria experienced the worst outbreak due to community transmission. At this time, vaccination was not available, and strict stay at home orders were the primary infection prevention and control measure used to reduce the spread of COVID-19.

In this section, we demonstrate the insights the SEIR-e model can provide into different exposure pathways by applying the model to COVID-19 case data, and estimating parameters which reflect transmission due to people–people and people–pathogen interactions. COVID-19 case data represents outbreaks where transmission occurred in an range of indoor environments such as households, workplaces and supermarkets, as well as outdoor environments. As the SEIR-e model is designed for specific indoor environments, it is applicable to a subset of the case data which is not readily available. Thus, we assume the population is located indoors for the duration of the outbreak to demonstrate the utility of the model, and acknowledge this assumption is a limitation of the modelling and results provided in this section.

### 3.1 Data

Case data relating to SARS-CoV-2 is available via public data repositories. We extract case data for Victoria from [36], and Fig 3(a) shows two distinct infectious peaks in 2020. The first peak can be attributed to overseas arrivals, so we isolate and study the second peak from 14/06/20–06/11/20 where the majority of cases were locally acquired (Fig 3(b)). A discontinuity in the number of infected people is evident during August, where the number of active cases drops from 7155 to 4864 to: “account for cases cleared over the past five days, supported by additional capacity”, [37]. Thus, this discontinuity is a result of public health record keeping, and a smooth dataset is produced using the 5 day moving average (Fig 3(b)).

**Figure 3:**
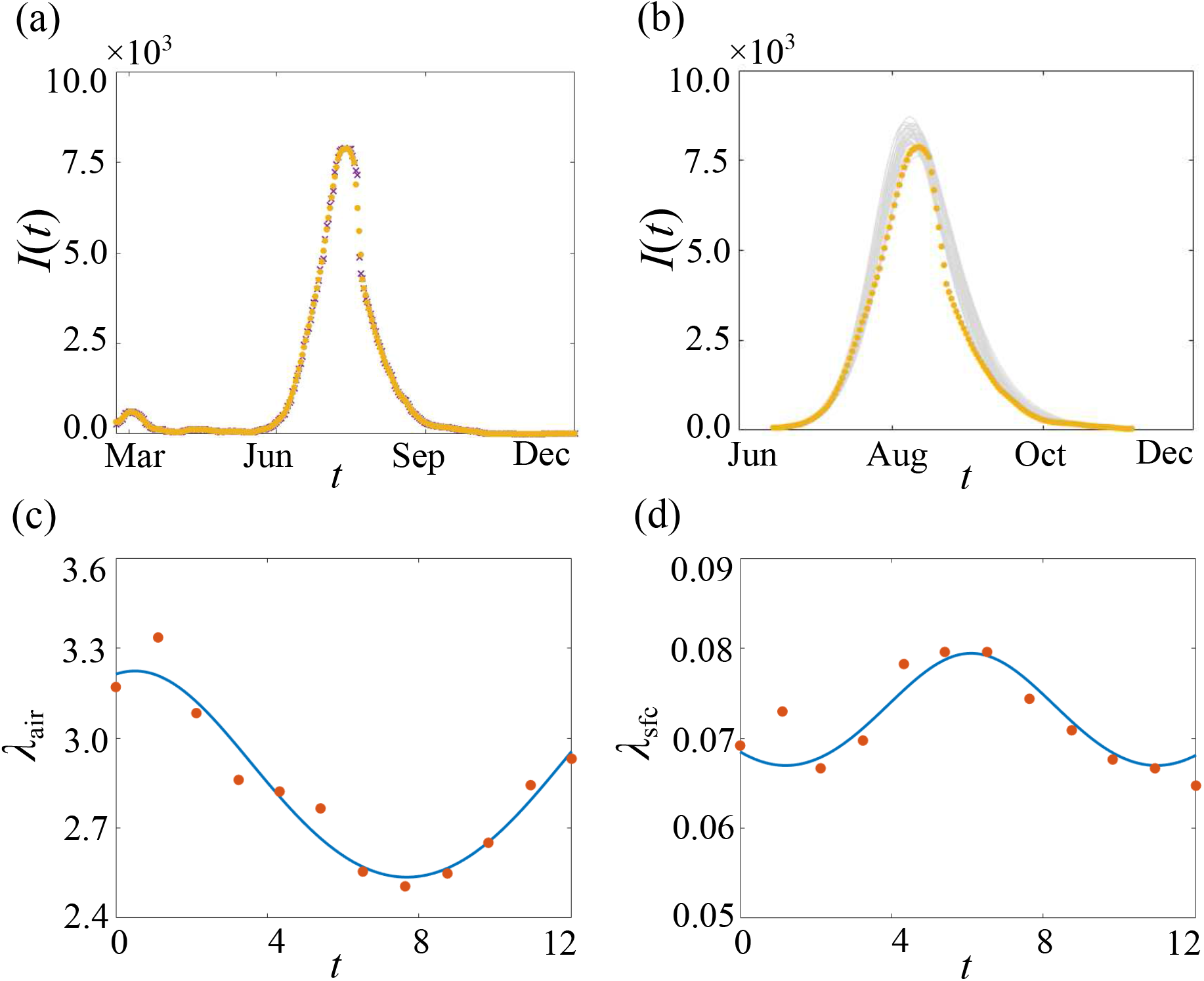
Victorian outbreak of COVID-19 in 2020. In (a), the 5 day moving average (yellow) is compared to raw case data (purple) [36], and to 50 randomly accepted parameter combinations (grey) in (b), where *I* is measured in thousands of people and *t* in months. Comparison of decay rates (dots) computed using historical weather observations [32–34] and fitted time dependent decay rates (line) for (c) airborne and (d) surface decay of COVID-19, where *λ*_air,sfc_ is measured in hours^−1^ and *t* in months.

### 3.2 Mathematical methods

Bayesian inference is a common parameter estimation technique which uses Bayes’ theorem, *π*(*θ*|*x*) ∝ *f* (*x*|*θ*)*π*(*θ*), where *π*(*θ*|*x*) is the posterior, *π*(*θ*) is the prior and *f* (*x*|*θ*) is the likelihood. The posterior distribution captures the relationship between the parameter *θ* and the observed data, *x*, while the prior distribution represents prior knowledge about appropriate values for *θ*. The likelihood represents a belief about *x* given *θ*, and is often intractable and costly to compute [38, 39]. Approximate Bayesian computation (ABC) methods are a class of likelihood free methods where a simulation– based process replaces the computation of the likelihood [38–40]. ABC methods are comprehensively reviewed in [38–40], so we briefly summarise the key points below.

ABC methods can be used to compare simulated data, *x*^*\**^, to the observed data, *x*. The simplest ABC algorithm is a rejection sampler where a candidate parameter, *θ*^*\**^, is sampled from *π*(*θ*), and accepted if the discrepancy between *x* and *x*^*\**^ is less than the required tolerance, *d*(*x, x*^*\**^) ≤*ϵ*. The Euclidean distance is commonly used as the discrepancy metric, *d*(*x, x*^*\**^) = ||*x* − *x*^*\**^||_2_. ABC methods result in a sample of parameters from *π*(*θ*|*d*(*x, x*^*\**^) ≤ *ϵ*) which is considered a good approximation of *π*(*θ*|*x*) if *ϵ* is sufficiently small [38–40]. We use ABC to infer posterior distributions for parameters of the SEIR-e model which correspond to disease transmission, people–people and people–pathogen interactions.

#### 3.2.1 Model equations

In the SEIR-e model, exposure to pathogens in the environment and the probability of infection is related using a dose–response function. Dose–response functions are reviewed in [23]. For simplicity, we use a linear dose–response function to determine the probability of infection due to inhalation and surface pickup. As the Victorian outbreak occurred over several months, time dependent decay rates are used to account for the effects of seasonal change on airborne and surface decay of SARS-CoV-2 (Fig 3(c,d), Appendix C.1). Thus, the model equations for the SEIR-e model when applied to the Victorian outbreak are:

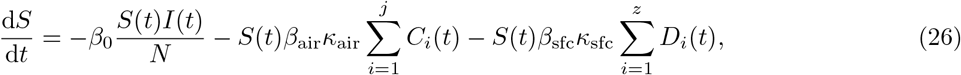

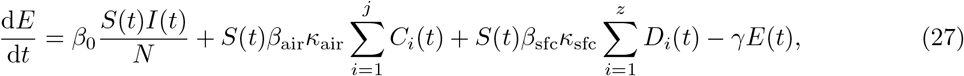

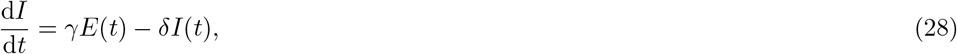

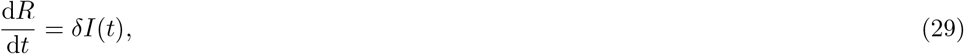

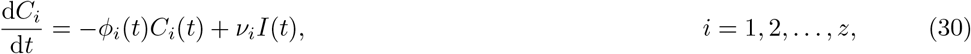

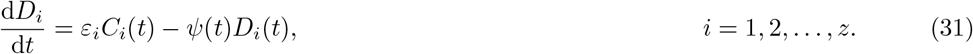

In constructing the model and applying it to the Victorian outbreak, we assume the contact parameter, *β*_0_, is constant. Realistically the value of *β*_0_ varied during the Victorian outbreak as strict stay at home orders and increased awareness limited transmission due to social interactions [41]. While a time dependent contact parameter could capture the effect of dynamic intervention measures on transmission and may be more realistic in the context of the Victorian outbreak, we emphasise the intent of this study is to demonstrate how a physical model can interface with a disease transmission model, and not to assess the effectiveness of infection prevention and control measures on controlling disease outbreaks. We acknowledge this is a limitation of the model and results provided, and suggest it for future consideration.

#### 3.2.2 Prior knowledge

To infer posterior distributions, we non-dimensionalise the model and use prior distributions which align with parameter values reported in the literature (Appendix C.2). Sources report values for the transmission rate ranging from 0.1–0.3, mean incubation periods of 3.6–7 days, and a median infectious period of 9.5 days [3, 35]. While values for *β*_air,sfc_ are not readily available, numerical exploration of the SEIR-e model reveals that values of 𝒪(10^−7^) are appropriate. Thus, we establish conservative uniform priors *β*_0_ ∼ *U* (0.1, 0.4), *β*_air,sfc_ ∼ *U* (0.5 × 10^−7^, 5 × 10^−7^), *γ*^−1^ ∼ *U* (3, 6), *δ*^−1^ ∼ *U* (9, 12) to fit the model parameters. We use the dataset to set *I*(0) = 40, *E*(0) = 65, *R*(0) = 0, *S*(0) = *N* − *E*(0) − *I*(0) − *R*(0) with *N* = 35 × 10^3^.

### 3.3 Results

To establish a baseline for comparison, we first infer parameters of the standard SEIR model, *θ*^†^ = (*β, γ*^−1^, *δ*^−1^), where 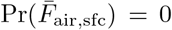, and find the posterior modes align with reported values, 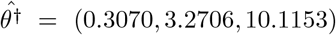 (Fig 4(a)–(c)). We infer parameters of the SEIR-e model, *θ* = (*β*_0_, *γ*^−1^, *δ*^−1^, *β*_air_, *β*_sfc_), and compute the posterior modes, 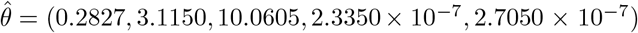 (Fig 4). As expected, the estimates of *γ*^−1^ and *δ*^−1^ are consistent between the two models and align with reported values. The SEIR-e model explicitly distinguishes between exposure due to people–people and people–pathogen interactions, while the standard SEIR model implicitly includes transmission due to different exposure pathways. Thus, it is reasonable that estimates of *β* are greater than *β*_0_, and comparing these estimates provides insight into the proportion of *β* which can be attributed to different exposure pathways. The similarities between the posteriors for *β*_air,sfc_ could imply that the contribution of inhalation and surface pickup on exposure are analogous, as implied in the phase plane analysis of the environment model (refer to Application to SARS-CoV-2).

**Figure 4:**
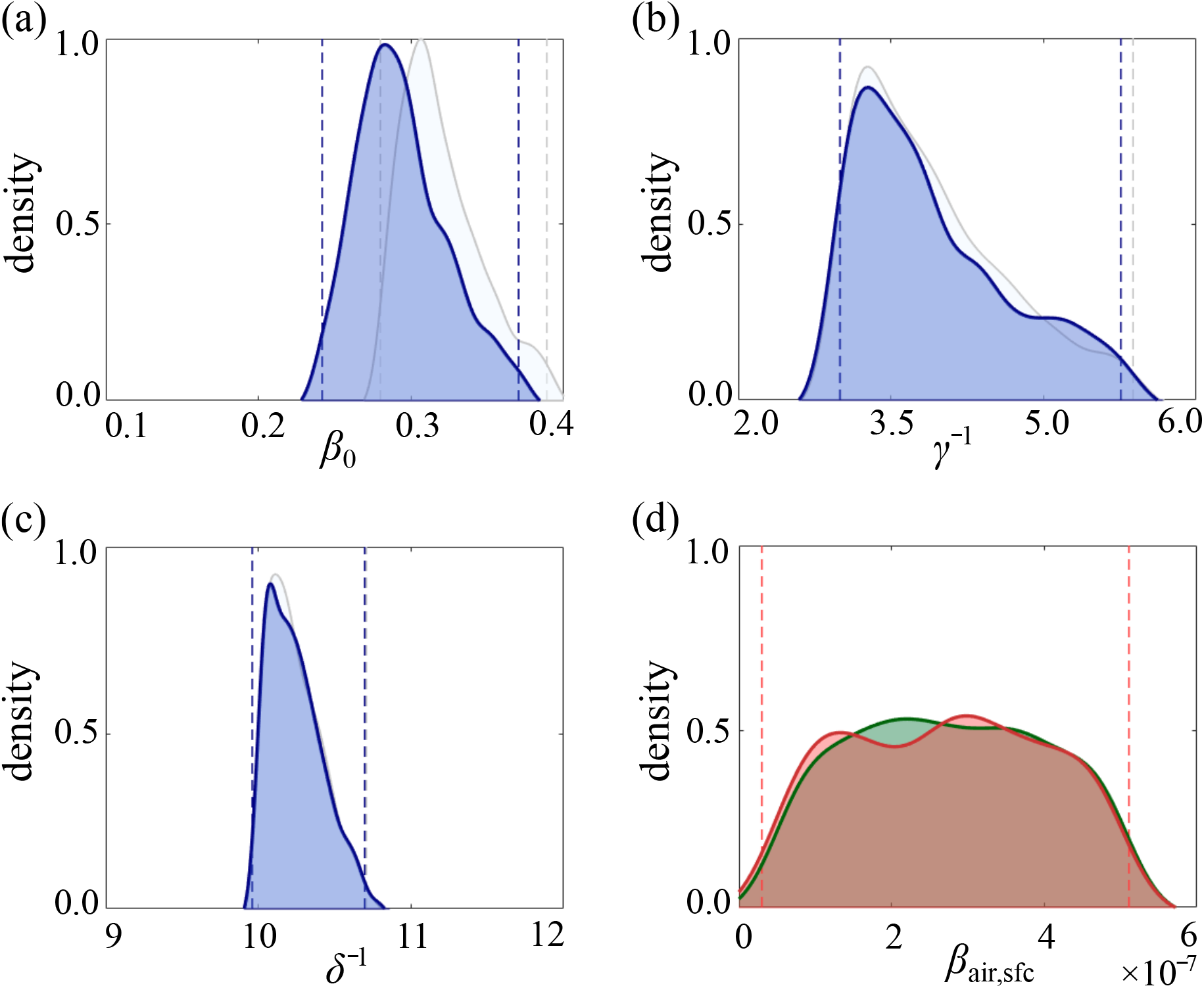
Comparison of the SEIR-e and SEIR models. In (a)–(c), the marginal posteriors of the SEIR-e model (blue) are compared to a standard SEIR model (grey), where the 95% credible interval is shown as the dashed lines. In (d), the marginal posterior for *β*_air_ (green) is overlaid with *β*_sfc_ (red), where the difference between the 95% credible intervals is not visually significant.

Like any virus, SARS-CoV-2 evolves over time and the properties of new variants, such as transmissibility, may differ between variants [42]. The 2020 Victorian outbreak of COVID-19 involved multiple variants which are no longer monitored due to little on going circulation [43]. The World Health Organisation has classified 5 variants of concern and 2 variants of interest for SARS-CoV-2 as of February 3, 2022 [42]. As the dataset used in this study does not involve current variants of concern or interest, the results in Fig 4 may not be valid when considering more recent outbreaks of the SARS-CoV-2 Delta or Omicron variants.

## 4 Conclusion

In this study, we present a novel SEIR-e model for disease transmission and its application to SARS-CoV-2. A physics–based environment model is developed to investigate how exposure to pathogens via inhalation and surface pickup results in infection. While the environment model is parametrised for SARS-CoV-2, the general modelling framework is applicable to other chemical, biological or radiological materials. Derivation of analytic solutions and linear stability analysis indicates that while viral deposition is the dominant decay process, the contribution of airborne and deposited material to the total dose received is equivalent in the long time limit. By coupling the environment model with a standard compartment model for disease transmission, we formulate the SEIR-e model to investigate the spread of infectious diseases due to people–people and people– pathogen interactions.

A key feature of the SEIR-e model is the distinction between exposure due to people–people and people–pathogen interactions, which is not explicitly included in standard disease transmission models [25–27]. This results in a expression for ℛ_0_(*t*) which accounts of people–people interactions, inhalation of viral aerosols, and fomite transmission. Another novelty is the coupling between indoor air quality and disease transmission, which captures the contribution of physical processes such as air exchange, viral decay and deposition on disease transmission. One advantage of using the SEIR-e model to study infectious diseases is that we gain insight into what proportion of the traditional contact parameter, *β*, accounts for people–people and people–pathogen interactions. A case study of the 2020 Victorian outbreak of COVID-19 demonstrates this where parameter values which describe people–people interactions differ significantly from *β*. Similar insight could be gained for other influenza like illnesses, and could be used to assess the impact of infection prevention and controls measures on reducing transmission.

There are many possible extensions of the environment and SEIR-e models developed in this study. In constructing the environment model, we take the most fundamental approach and use simplistic mechanisms to describe physical processes. One extension is to incorporate more complex processes which better describe the physical system [13]. As the environment model is not spatially resolved, another extension is to include a spatial dimension by dividing the environment into multiple spatial compartments [44]. This extension may require additional information or assumptions about the transport of airborne aerosols in indoor environments. As previously discussed, a limitation of the SEIR-e model is the time independent contact parameter, *β*_0_. Incorporating a time dependent contact parameter may account for changes in social and behavioural patterns as a result of dynamic infection prevention and control measures, and the corresponding effect on disease transmission [45]. Historically strict and dynamic intervention methods are not often associated with disease outbreaks.

Thus, the model remains relevant in the majority of contexts, and we leave the question of incorporating a time dependent contact parameter for future consideration. Further extensions of the SEIR-e model include using additional compartments to distinguish between individuals at different stages of infection, or between those who are symptomatic and asymptomatic [3, 18]. As COVID-19 is a new and rapidly evolving disease, other extensions include adapting the SEIR-e model to account for different variants of SARS-CoV-2 [46], the possibility of mutation [47], and the effect of vaccination [48, 49] on transmission.

## Data Availability

MATLAB implementations of key numerical algorithms are available on GitHub.

https://github.com/tamaratambyah/Tambyah2022

## Data access

MATLAB implementations of key numerical algorithms are available on GitHub.

## Author Contributions

All authors contributed equally to the design of the study. TAT performed numerical simulations and drafted the article. All authors gave approval for publication.

## Ethics

This study did not require ethics approvals.

## Acknowledgements

We appreciate the helpful comments provided by our colleagues Dr Ralph Gailis and Dr Peter Dawson.

## A Environment model

In this section, we provide additional details for the environment model, and results to investigate the sensitivity of the model to different parameters. The environment model is restated here for convenience:

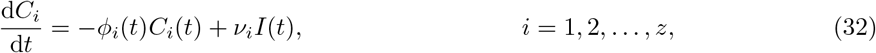

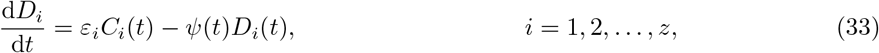

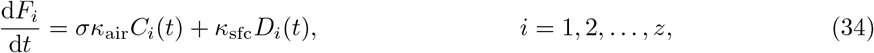

where

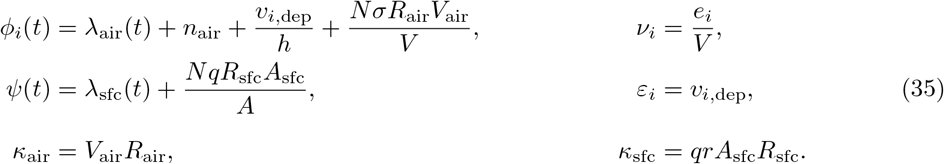

### A.1 Non-dimensionalisation

To non-dimensionalise Eqs (32)–(34), we choose

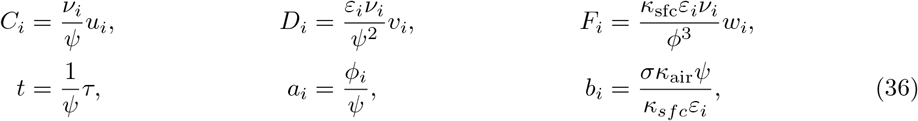

where *u*_*i*_, *v*_*i*_, *w*_*i*_ and *τ* are non-dimensional. Substituting Eq (36) into Eqs (32)–(34) gives the non-dimensional model as:

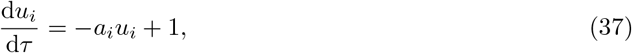

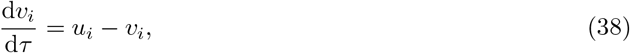

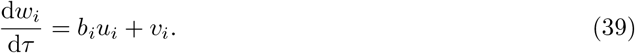

### A.2 Sensitivity analysis

We consider the scenario with *I*(*t*) = 1 and *N* = 2 (refer to Application to SARS-CoV-2 in the main document), and conduct parameter sweeps to investigate the sensitivity of the environment model to parameter values relating to SARS-CoV-2. We fix the size and environmental conditions of the indoor environment (Table 1), and choose to vary a selection of dimensional parameters in Eq (35). As some parameters depend on or are directly influenced by other parameters, two related parameters are varied simultaneously, while all other parameters remain fixed at the values given in Table 1. For each parameter combination, we compare the probability of infection after 8 hours of exposure to determine which parameter combinations yield the highest probability of infection.

**Table 1:** Environment model default parameter values.

|  | Symbol | Description | Value (units) |
| --- | --- | --- | --- |
| Fixed | $V$ | Room volume | 108 (m<sup>3</sup>) |
| | $h$ | Room height | 3 (m) |
| | $A$ | Horizontal surface area | 10 (m<sup>2</sup>) |
| | $\lambda_{\text{air}}$ | Airborne decay rate at 40% RH and 23 °C | 0.6 (hr<sup>-1</sup>) |
| | $\lambda_{\text{sfc}}$ | Surface decay rate at 40% RH and 23 °C | 0.06 (hr<sup>-1</sup>) |
| Varied | $n_{\text{air}}$ | Air exchange rate | 2 (changes hr<sup>-1</sup>) |
| | $j$ | Number of size bins in respirable range | 30 (bins) |
| | $\kappa_{\text{air}} = V_{\text{air}} R_{\text{air}}$ | Breathing rate | 10 (L min<sup>-1</sup>) |
| | $R_{\text{sfc}}$ | Surface contact rate | 5 (contacts hr<sup>-1</sup>) |
| | $A_{\text{sfc}}$ | Surface contact area | 0.0128 (m<sup>2</sup>) |
| | $q$ | Pickup efficiency | 0.1 |
| | $r$ | Transfer efficiency | 0.34 |
| | $N_{\text{VL}}$ | Viral load | $7 \times 10^6$ (virus mL<sup>-1</sup>) |
| | $R_{\text{c}}$ | Coughing rate | 1 (cough min<sup>-1</sup>) |
| | $\rho$ | Probit slope of dose-response | 3.6 |
|  | ID<sub>50</sub> | Dose for 50% probability of infection | 189 (virus) |

#### A.2.1 Contact and inhalation parameters

The total dose is comprised of an inhalation and contact component. Eq (34) shows that the total dose, and thus the probability of infection, is linearly proportional to each component and to variation in *κ*_air,sfc_. However, the inhalation component of the total dose is constrained by droplet size, as only droplets in the respirable range, 1 ≤ *i* ≤ *j*, are inhaled. Another assumption is that the environment is well–ventilated such that the airborne contaminant exchanges with ambient air at a rate *n*_air_. To investigate the effect of these assumptions, we vary *n*_air_ and *j* simultaneously. The settling time for droplets with diameter 250 *μ*m is approximately 6 seconds. We assume that 6 seconds is sufficient for droplets to be inhaled at least 1.5 m from the source, and choose *j* values ranging from 30–250 bins, and *n*_air_ values ranging from 1–5 changes hr^−1^.

As the contact component is not constrained by droplet size, equivalent changes in the dimensional parameters which form *κ*_sfc_ will yield equivalent sensitivity. For example, increasing *A*_sfc_ by a factor of 2 will have the same effect as increasing *R*_sfc_ by a factor of 2. However, as each parameter is bound by appropriate values found in the literature, we vary parameters within reasonable limits to determine which parameters have a greater influence on the probability of infection. Methods for measuring the surface area of the hand vary between studies, resulting in highly diverse values for *A*_sfc_ which depend on biological factors such as age, gender and race [50–52]. As SARS-CoV-2 was mainly transmitted between adults at the time of the Victorian outbreak, we compare *A*_sfc_ values for adults of different races. A study which considers mainly Caucasian adults reported that *A*_sfc_ = 0.0128 m^2^ [52], whereas a study which considers Asian adults found that *A*_sfc_ = 0.0139 m^2^ [51]. One study that considers a mix of races reported that *A*_sfc_ = 0.027 m^2^ [50]. Thus, we compare *A*_sfc_ values ranging from 0.0128–0.027 m^2^, and we choose *R*_sfc_ values ranging from 1–13 contacts hr^−1^. Appropriate values for *q* and *r* vary depending on the type of contaminated material or infection being considered [53]. As no average value for *q* and *r* is available for SARS-CoV-2, we vary *q* and *r* from 0.1–0.3.

Fig 5 demonstrates that the model is significantly sensitive to variation in *q* and *r*, sensitive to *A*_sfc_ when 3 *< R*_sfc_ *<* 9 contacts hr^−1^, and not sensitive to variation in *n*_air_ or *j*. Thus, we conclude that variation in the parameters used to compute the contact component of the total dose will have a greater effect on the probability of infection than the parameters used to compute the inhaled component. It is important to note that the results presented in Fig 5 and the corresponding conclusions drawn relate to a particular set of parameters for the BLO model and dose response curve (Eq (9)) given in Table 1. If the value of these parameters change, the conclusions stated above may no longer be valid.

**Figure 5:**
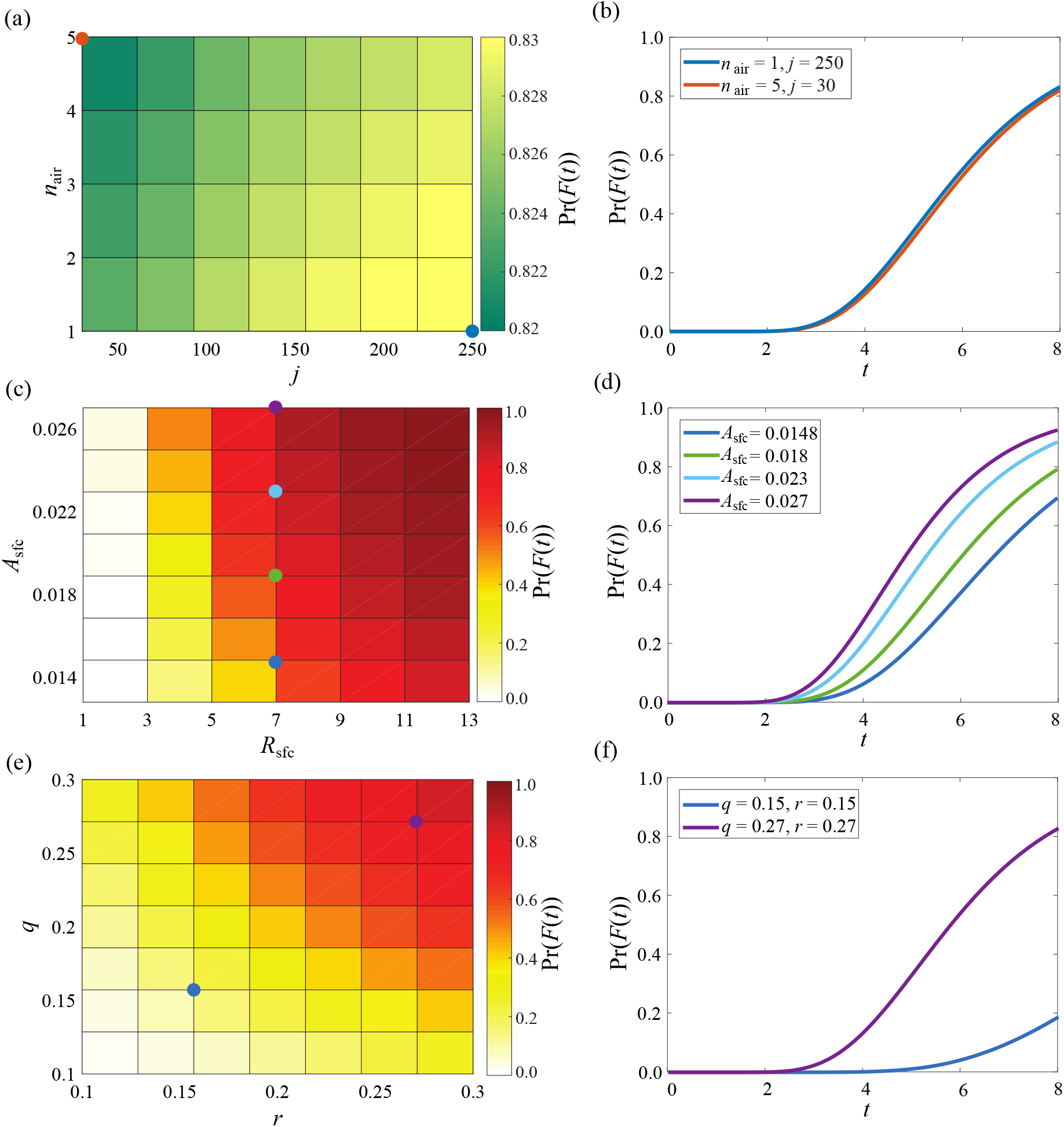
Sensitivity of the environment model to contact and inhalational parameters for SARS-CoV-2. In (a,c,e), parameters are varied simultaneously where the colour represents the probability of infection after 8 hours of exposure. The coloured dots correspond to the parameter combinations shown in (b,d,f).

### A.2.2 Viral parameters

In the context of SARS-CoV-2, a BLO model is used to determine the emission rate of viral droplets in the *i*^th^ size bin, *e*_*i*_, due to an infected individual coughing, speaking and breathing [31]. While other source terms such as sprayers or nebulisers could also be represented, the BLO model and its application to SARS-CoV-2 is detailed in [31]. A number of viral parameters govern *e*_*i*_, including the coughing rate, *R*_c_, the breathing rate, *κ*_air_, and the viral load, *N*_VL_. Values for *R*_c_ vary between individuals and infections, with no average value available for SARS-CoV-2, and values for *κ*_air_ vary depending on body weight, age and activity levels. One study reports that the maximum resting breathing rate for an adult (older than 12 years of age) is 0.137 L/minute per kilogram of body weight, whereas the maximum moderate breathing rate is 0.384 L/minute per kilogram of body weight [54]. For high intensity activities, such as running, the maximum breathing rate is 0.962 L/minute per kilogram of body weight [54]. As we are considering environments such as workplaces and residential buildings, we choose breathing rate values ranging from 0.137–0.384 L/minute per kilogram of body weight, and assume an average adult weighs 80 kg. Fig 6(a,b) demonstrates that the probability of infection increases as *κ*_air_ increases for small coughing rates, *R*_c_ ≤ 1 coughs min^−1^. This is expected given the linear relationship between *κ*_air_ and the total dose (Eq (34)).

**Figure 6:**
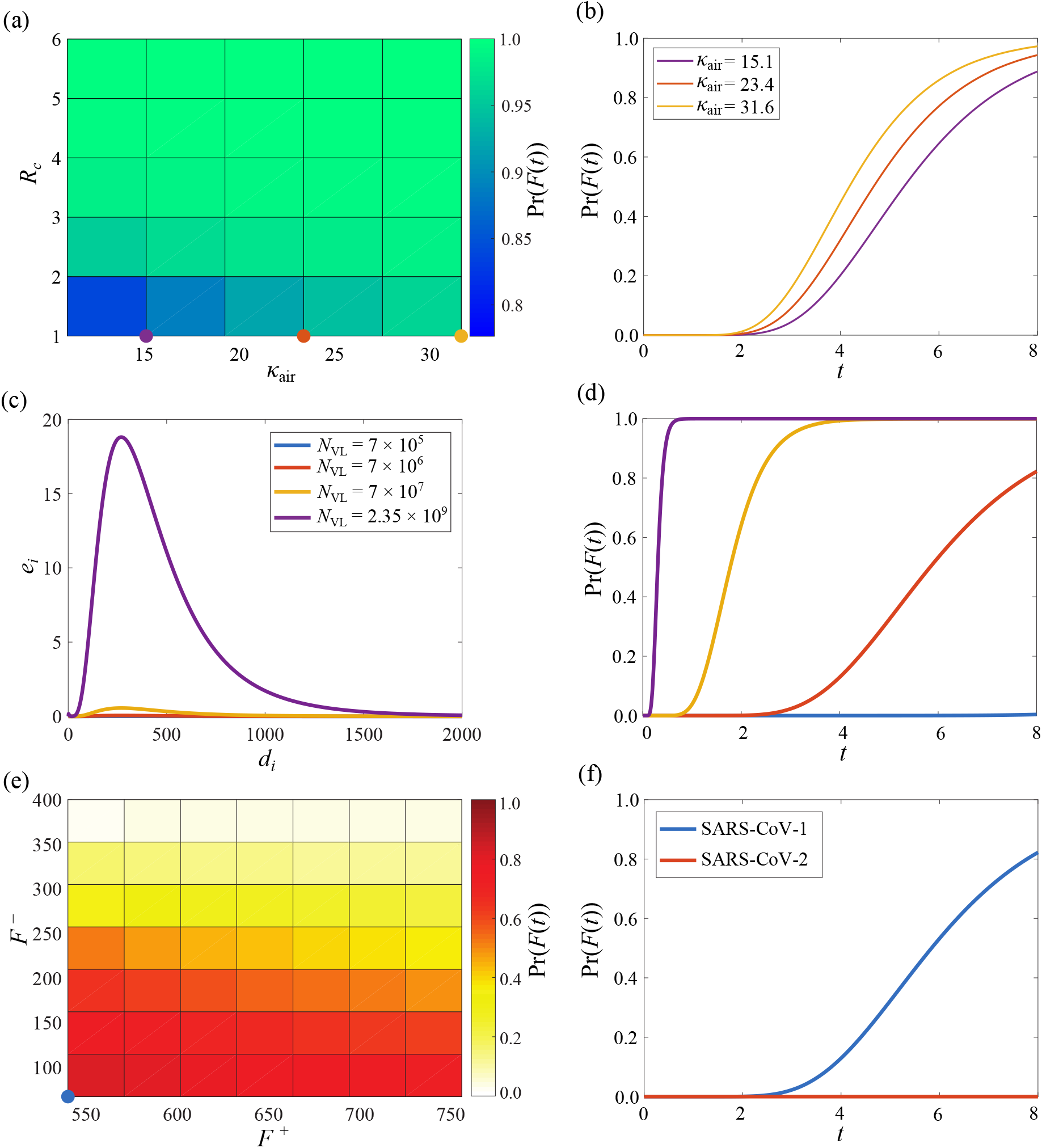
Sensitivity of the environment model to viral parameters for SARS-CoV-2. In (a,e), parameters are varied simultaneously where the colour represents the probability of infection after 8 hours of exposure. The coloured dots correspond to the parameter combinations shown in (b,f). In (c,d), viral emission and corresponding probability of infection curves are shown for different *N*_VL_ values.

The viral load, *N*_VL_, is a measure of the total number of viral particles present per unit volume, and can vary between different samples, such as nasopharyngeal and oropharyngeal swabs and sputum samples. One study found that the average viral load for SARS-CoV-2 in nasopharyngeal and oropharyngeal swabs is 6.76 × 10^5^ virus mL^−1^ with a maximum value of 7.11 × 10^8^ virus mL^−1^ [55]. This study also reported that the average viral load in sputum samples is 7 × 10^6^ virus mL^−1^ with a maximum value of 2.35 × 10^9^ virus mL^−1^ [55]. As reported *N*_VL_ values vary between orders of magnitude, the model is significantly sensitive to *N*_VL_ (Fig 6(c,d)).

The dose–response curve (Eq (9)) used to compute the probability of infection in Figs 2(c), 5 and 6(a)–(d) is parametrised by ID_50_ = 189 virus and *ρ* = 3.6. These values are based on a study of SARS-CoV-1 in mice which recorded an infectious dose of *F* (*t*) ∈ [67, 540] plaque forming units (PFU) [56], and were used in the original study [31] as no experimental data for SARS-CoV-2 was available at the time of publication. A recent study of SARS-CoV-2 in mice recorded an infectious dose of *F* (*t*) ∈ [504, 756] PFU [57], which corresponds to ID_50_ = 617.3 virus and *ρ* = 18.7. The infectious dose, *F* (*t*) ∈ [*F* ^−^, *F* ^+^], and values for ID_50_ and *ρ* vary greatly between the two studies as they relate to different pathogens, and potentially different types of genetically modified mice. Until more experimental data relating to SARS-CoV-2 is made available, appropriate values for *F* ^−^ and *F* ^+^ can only be informed by current data [57], data relating to other CoVs [56], and numerical exploration.

Assuming Pr (*F* (*t*) = *F* ^−^) = 0.05 and Pr (*F* (*t*) = *F* ^+^) = 0.95, we compute

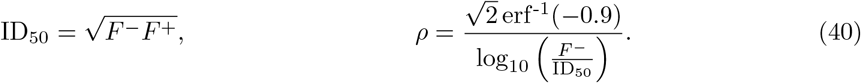

Fig 6(e,f) compares the probability of infection when the dose–response curve is parametrised by SARS-CoV-1 and SARS-CoV-2 for an exposure period of 8 hours. If the exposure duration changes, the results and conclusions drawn may no longer be valid.

## B Basic reproduction number

The basic reproduction number is the spectral radius of the Next Generation matrix, *K* = *FV*^−1^, where 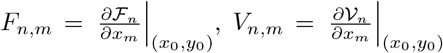 and (*x*_0_, *y*_0_) is the disease free equilibrium (DFE) [23,24,30]. The DFE is determined by solving Eqs (17)–(22) equal to zero. This results in *I* = *E* = 0, which implies *C*_*i*_ = *D*_*i*_ = 0 ∀*i* and 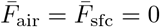. Thus, for a fully susceptible population the DFE is (*S*_0_, *E*_0_, *I*_0_, *R*_0_, *C*_*i*,0_, *D*_*i*,0_)^*T*^ = (*N*, 0, 0, 0, 0, 0)^*T*^.

As only the first entry of ℱ is non-zero, *F* only has non-zero entries in the first row. This implies that *K* will only have non-zero elements in the first row, such that 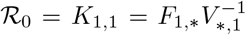, where *F*_1,*\**_ and 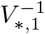 represent the first row and column of *F* and *V*^−1^ respectively. We find that

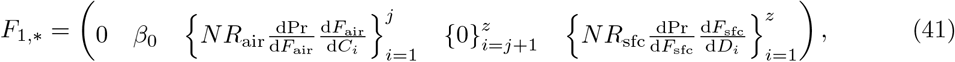

and that *V* is a lower triagonal matrix consisting mostly of zeros, with non-zero elements

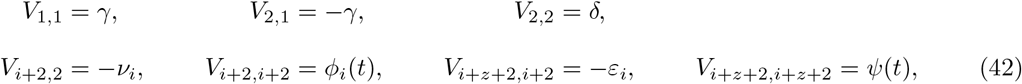

for *i* = 1, 2, …, *z*. For example, if *z* = 2

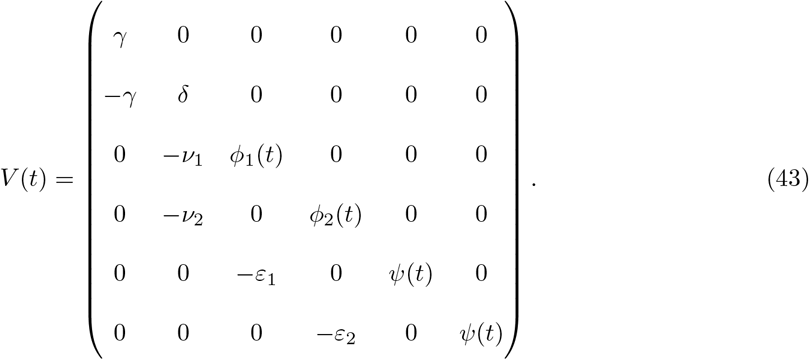

Using Gaussian elimination, we compute

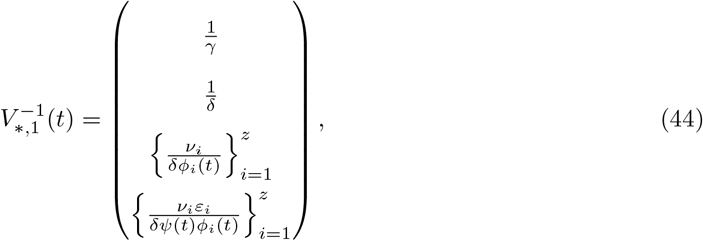

such that

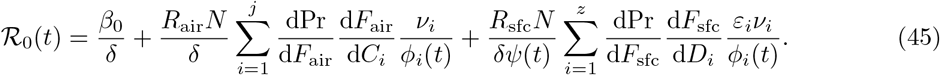

## C COVID-19 case study

### C.1 Time dependent decay rates

To compute time dependent decay rates for SARS-CoV-2, we use monthly historical data for the 3pm temperature (*T*) and relative humidity (RH) in Melbourne [34] (Table 2). As the *T* and RH values correspond to outdoor weather conditions, we use a UV index of 1 to extract half life values for airborne 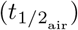 and surface 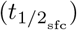 decay from the US Department of Homeland Security [32, 33]. Thus, we subsequently assume that outdoor environmental conditions are replicated indoors. Where no half life value is available for exact *T* and RH conditions, the closest value is used. As *λ* = ln 2*/t*_1*/*2_ for exponential decay models, we compute monthly decay rates to which we fit a periodic curve (Fig 3(c,d)).

**Table 2:** Half life values for airborne and surface decay of SARS-CoV-2 for historical weather conditions in Melbourne.

|  | Month |  |  |  |  |  |  |  |  |  |  |  |
| --- | --- | --- | --- | --- | --- | --- | --- | --- | --- | --- | --- | --- |
|  | Jan | Feb | Mar | Apr | May | Jun | Jul | Aug | Sep | Oct | Nov | Dec |
| $T$<br>(°C) | 24.2 | 24.7 | 22.8 | 19.6 | 16.3 | 13.7 | 13.0 | 14.3 | 16.1 | 18.3 | 20.4 | 22.4 |
| RH<br>(%) | 47 | 48 | 49 | 52 | 59 | 63 | 61 | 56 | 53 | 50 | 49 | 47 |
| $t_{1/2_{\text{air}}}$<br>(min) | 13.12 | 12.47 | 13.49 | 14.54 | 14.74 | 15.04 | 16.28 | 16.6 | 16.32 | 15.69 | 14.63 | 14.19 |
| $t_{1/2_{\text{sfc}}}$<br>(hr) | 10.02 | 9.5 | 10.4 | 9.94 | 8.86 | 8.71 | 8.71 | 9.32 | 9.78 | 10.25 | 10.4 | 0.71 |

### C.2 Non-dimensionalisation

SEIR models for disease transmission are uni-directional. That is, the entire population is considered susceptible with the underlying assumption that the majority of susceptible individuals become infected. This assumption is not realistic when modelling COVID-19 as due to infection prevention and control measures such as stay at home orders, social distancing and increased hand hygiene, only a small fraction of susceptible people become infected. As the Victorian outbreak resulted in approximately 0.3% of the Victorian population being infected, we distinguish between the expected number of people susceptible to infection, *N*, and the total population of Victoria, *N*_*T*_, through non-dimensionalisation. In this study, *N* = 35 × 10^3^ and *N*_*T*_ = 6.28 × 10^6^.

To non-dimensionalise Eqs (26)–(31), we choose

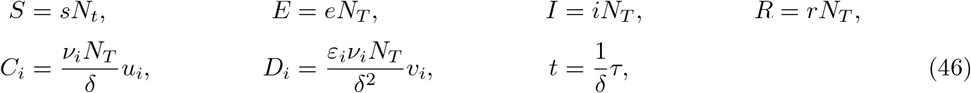

such that *s, e, i, r, u*_*i*_, *v*_*i*_ and *τ* are non-dimensional. Substituting Eq (46) into Eqs (26)–(31) gives the non-dimensional model:

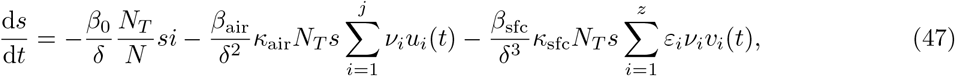

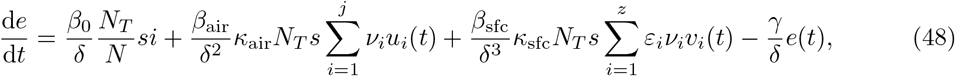

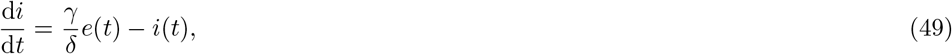

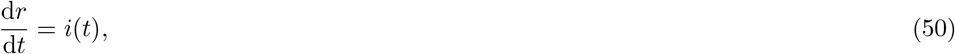

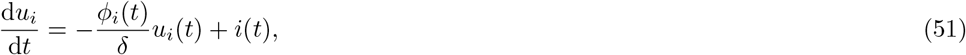

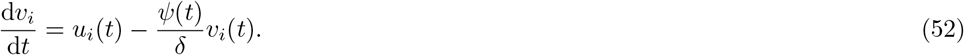

for *i* = 1, 2, …, *z*. Non-dimensionalising the model in this way scales the people–people interaction term by *N*_*T*_ */N* in order to account for the large difference between *N* ≈ *S*(0) and *N*_*T*_.

